# Analyzing Supply and Demand on a General Internal Medicine Ward: A Cross-Sectional Study

**DOI:** 10.1101/2021.05.20.21257349

**Authors:** Michael Fralick, Neal Kaw, Mingkun Wang, Muhammad Mamdani, Ophyr Mourad

## Abstract

**Background:** The capacity of the general internal medicine clinical teaching units has been strained by decreasing resident supply and increasing patient demand. The objective of our study was to quantitatively compare the number of residents (supply) with the volume and duration of patient care activities (demand) to identify inefficiency.

**Methods:** Using the most recently available data from an academic teaching hospital, we identified each occurrence of a set of patient care activities that took place on the clinical teaching unit. We completed a descriptive analysis of the frequencies of these activities and how the frequencies varied by hour, day, week, month, and year. Patient care activities included admissions, rounds, responding to pages, meeting with patients and their families, patient transfers, discharges, and responding to cardiac arrests. The estimated time to complete each task was based on the available data in our electronic healthcare system and interviews with general internal medicine physicians or trainees. To calculate resident utilization, the person-hours of patient care tasks was divided by the person-hours of resident supply. Resident utilization was computed for three scenarios corresponding to varying levels of resident absenteeism.

**Results:** Between 2015 and 2019 there were 14,581 consultations to general internal medicine from the emergency department. Patient volumes tended to be highest during January and lowest during May and June; and highest on Monday morning and lowest on Friday night. Daily admissions into hospital from the emergency department were higher on weekdays than on weekends, and hourly admissions peaked at 8:00 AM and between 3:00 PM and 1:00 AM. Weekday resident utilization was generally highest between 8:00 AM and 2:00 PM and lowest between 1:00 AM and 8:00 AM. In a scenario where all residents were present apart from those who were post-call, resident utilization generally never exceeded 100%; in scenarios where at least one resident was absent due to illness and/or vacation, it was common for resident utilization to approach or exceed 100%, particularly during daytime working hours.

**Interpretation:** Analyzing supply and demand on a general internal medicine ward has allowed us to identify periods where supply and demand are not aligned and to empirically demonstrate the vulnerability of current staffing models. These data have the potential to inform and optimize scheduling on an internal medicine ward.

## INTRODUCTION

As Competency By Design is being implemented for residency training programs across North America, fewer residents are rotating through the general internal medicine clinical teaching unit.^1,2^ Simultaneously, the complexity, acuity, and length of hospitalization for patients admitted to hospital has been increasing.^3^ The effect of decreasing resident supply and increasing patient demand has placed strains on the general internal medicine clinical teaching unit, leading some to question whether the current model requires redesign.^4^

Empirical studies quantifying the workload on a general internal medicine ward are either specific to individual tasks or lacking altogether. A prospective study at two general internal medicine wards quantified the time and tasks required to discharge a patient from hospital.^5^ They identified that the time to complete a the discharge paperwork was approximately 30 minutes, though additional time was required to fill out additional referrals and communicate the information to both patients and providers.^5^ A survey study of over 500 general internal medicine physicians in the United States identified that patient workload often exceeded the available staffing.^6^ While the physicians surveyed indicated that their workload likely affected patient safety, the study lacked data quantifying their daily tasks (i.e., demand) or the number of clinicians available to help with the tasks (i.e., supply). ^6^ The objective of our study was to model the supply (i.e., number of residents) and demand (i.e., patient care activities) on the general internal medicine clinical teaching unit at an academic teaching hospital to understand how changes in either variable could lead to mismatch, redundancy, and/or inefficiency.

## METHODS

### Study Setting

We conducted a modeling study based on a retrospective cohort of patients hospitalized under the general internal medicine service at St. Michael’s Hospital, an academic teaching hospital in Toronto, Ontario, using data up to 2019 (most recent available data). At St. Michael’s Hospital, there were approximately 80 inpatient general internal medicine beds cared for by five medical teams. Four of the five teams were each comprised of one staff physician, one senior resident (i.e., second-year resident or higher), three junior residents (i.e., one general internal medicine first-year resident and two non-general internal medicine residents), and two medical students. The fifth team was comprised of one staff physician and generally one senior resident. The number of teams and their breakdowns are consistent with the current St. Michael’s Hospital model.

In the current St. Michael’s Hospital model, one resident physician is in-hospital for four of the five teams for approximately 26 hours, and the remaining residents generally work from 8:00 AM until 5:00 PM Monday to Friday. On Saturday and Sunday, one resident is in hospital 24 hours per day for each team. On the fifth team, the resident works Monday to Friday from 8:00 AM until 5:00 PM. On the weekend this team is covered by the staff physician from 8:00 AM until 12:00 PM and covered by one of the in-house residents for the remaining hours.

### Data Sources

We modeled the current supply and demand on the general internal medicine ward. To model the demand, we identified the patient care tasks that occurred on the clinical teaching unit. They were identified through discussions with medical students, residents, and staff physicians. These tasks included: admitting patients into hospital from the emergency department, rounding on admitted patients, responding to pages, meeting with patients and their families, transferring patients out of the intensive care unit to the general internal medicine ward, discharging patients from hospital, and responding to cardiac arrests. For each task we retrieved historical data to understand the number of times each task occurred per day, the timing of the task to the nearest hour, and how the frequency of tasks varied by hour, day, week, month, and year. The exact dates for components generally spanned from 2015 to 2019, and some components had data spanning as far back as 2013 (Appendix 1).

Data for tasks related to patient care (i.e., admissions, daily rounding, and discharges) were accessed through our electronic medical record system, which automatically captures these data. Specifically, the electronic medical record has a log of all admissions, discharges, and the patient census for each day. Paging data were extracted directly from the hospital’s source paging system which captures the date and time of pages to each team’s pager. Cardiac arrest data were accessed through the hospital’s cardiac arrest database, which captures the date and time of cardiac arrests that occurred. Data related to the number of family meetings, medical consultations, and the time necessary to complete these tasks were estimated by interviewing staff physicians (N=3), internal medicine residents (N=3), and non-internal medicine residents (N=3). The s taff physicians were all general internal medicine physicians who regularly work on the inpatient GIM team. The medical students and residents were either currently working on GIM or had recently completed their GIM rotations (i.e., within the preceding 3 months).

### Statistical Analysis

For the year from February, 2018 to February, 2019, we calculated the duration of patient care tasks against available resident time. We divided each day into four periods whose boundaries either mark shifts in the number of working residents or shifts in the volume of aggregate patient demand (8:00 AM to 2:00 PM, 2:00 PM to 5:00 PM, 5:00 PM to 1:00 AM, and 1:00 AM to 8:00 AM). Within each period, we estimated total workload (demand) in units of person-hours by multiplying the frequency of each task by its estimated duration, and summing over all tasks. Within each period, we also estimated total supply. For example, if there were three residents available between 8:00 AM and 5:00 PM then there were 27 person-hours of resident time for patient care.

We then used the total demand and supply to estimate resident utilization in each period (Appendix 2). In particular, we divided the person-hours of demand by the person-hours of supply (i.e., resident time) to estimate the percentage of available resident time that was utilized in each period. We completed these computations for each period in one year of data, generating 52 data points to visualize the distribution of utilization in each period on each day of the week. We also performed a sensitivity analysis in which we assumed each patient task took 50% longer, to assess how a 50% increase in patient task load affected the supply and demand estimates.

The supply and demand were calculated in the current model under three scenarios. Scenario 1 assumed all residents were present apart from those who were post-call. Scenario 2 assumed one resident was away on vacation each week and also accounted for a resident being post-call. Scenario 3 assumed one resident was away on vacation and another resident was away sick, and also accounted for a resident being post-call. When the time of the consultation being requested was available, we utilized this time and no imputation was required. For patient records missing a consult request timestamp, we estimated the time the consult was requested by calculating the median time between consult request and consult completion/admission into hospital using data from patients where the consult request time was not missing. Specifically, we calculated the median time between consult completion/admission to hospital and the time of the consult request, and then subtracted this median time from the completion/admission time to impute the consult request time in instances where this particular timestamp was unavailable. All statistical analyses were performed using R version 3.6.2. This study was approved by the St. Michael’s Hospital Research Ethics Board.

## RESULTS

Between 2015 and 2019 there were 14,581 consultations to general internal medicine from the emergency department at St. Michael’s Hospital. Patient volumes generally tended to be highest during January and lowest during May and June (Figure 1A). Patient volumes also varied by day of week and hour of day, with the highest number of admitted patients typically occurring on Monday morning and the lowest number on Friday night (Figure 1B). Admissions into hospital from the emergency department varied by time of day with peaks observed at 8:00 AM and between 3:00 PM and 1:00 AM, and the weekdays generally had higher numbers of admissions per day compared to the weekend (Figure 1C, 1D). Similar patterns were observed for number of hospital discharges and number of pages to the on-call resident (Appendix 2). Cardiac arrests occurred more often on weekdays compared to weekends and were most likely to occur between 10:00 AM and 1:00 PM.

**Figure 1A.**
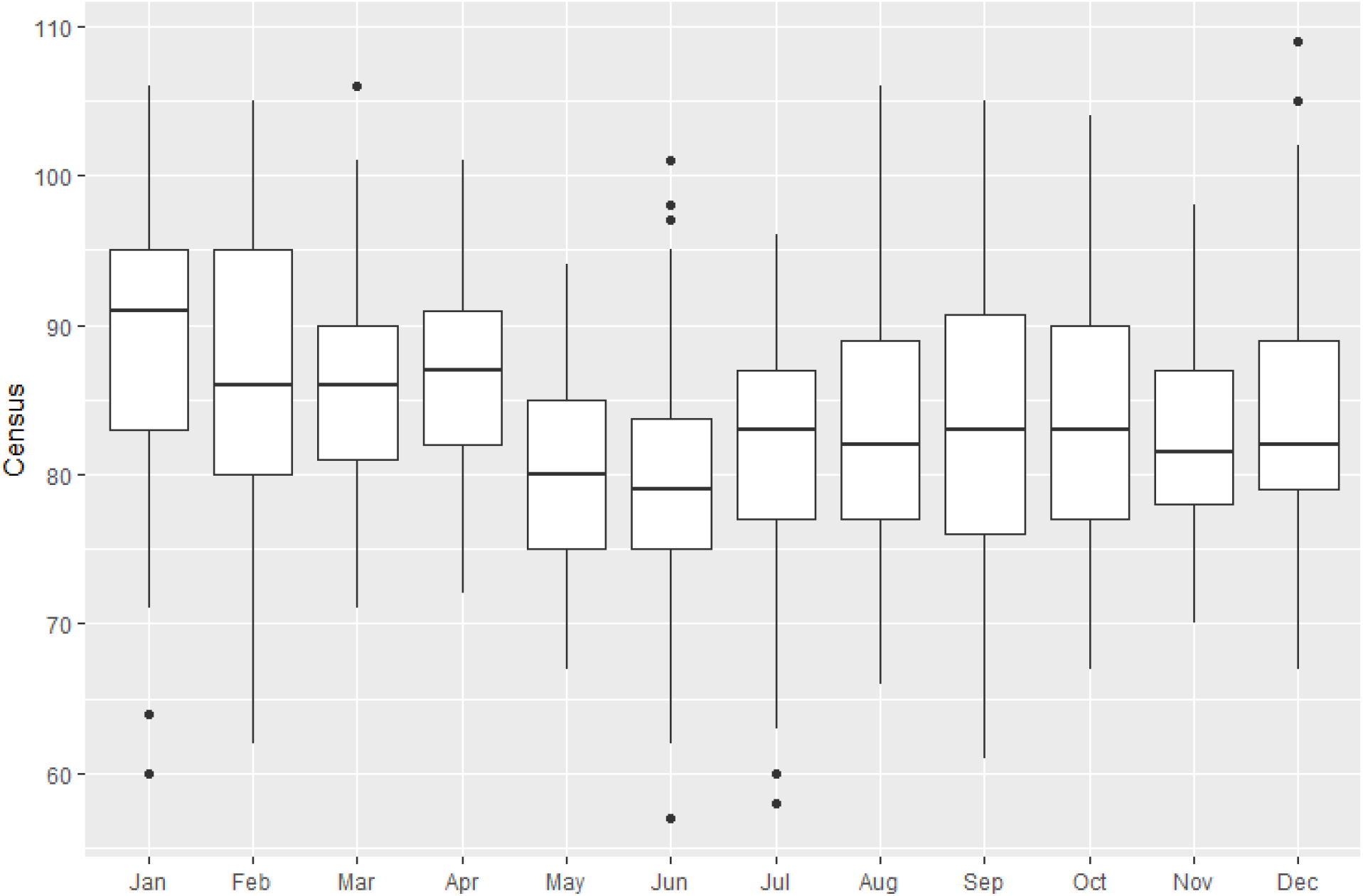
Distribution of 8:00 AM census in GIM inpatient service, by month Legend: Data from January, 2016 to December, 2018. For the census data, box-whisker plot is used to show an indication of how the census volumes are spread out. The lower and upper bounds of the box represent the 25^th^ percentile and the 75^th^ percentile of the volume of census, respectively, while the horizontal line inside the box represents the median of the data. The range that the box covers (25^th^ percentile to 75^th^ percentile) is defined as interquartile range (IQR), and the lower and upper ends of the whiskers represent the “minimum” (i.e., 25^th^ percentile – 1.5*IQR) and “maximum” (i.e., 75^th^ percentile + 1.5*IQR) observation in the sample, respectively. The black dots are outliers. This plot shows that the census volumes are generally highest from January to April and that the highest variability in the census generally occurred in January, February, September, and October.

**Figure 1B.**
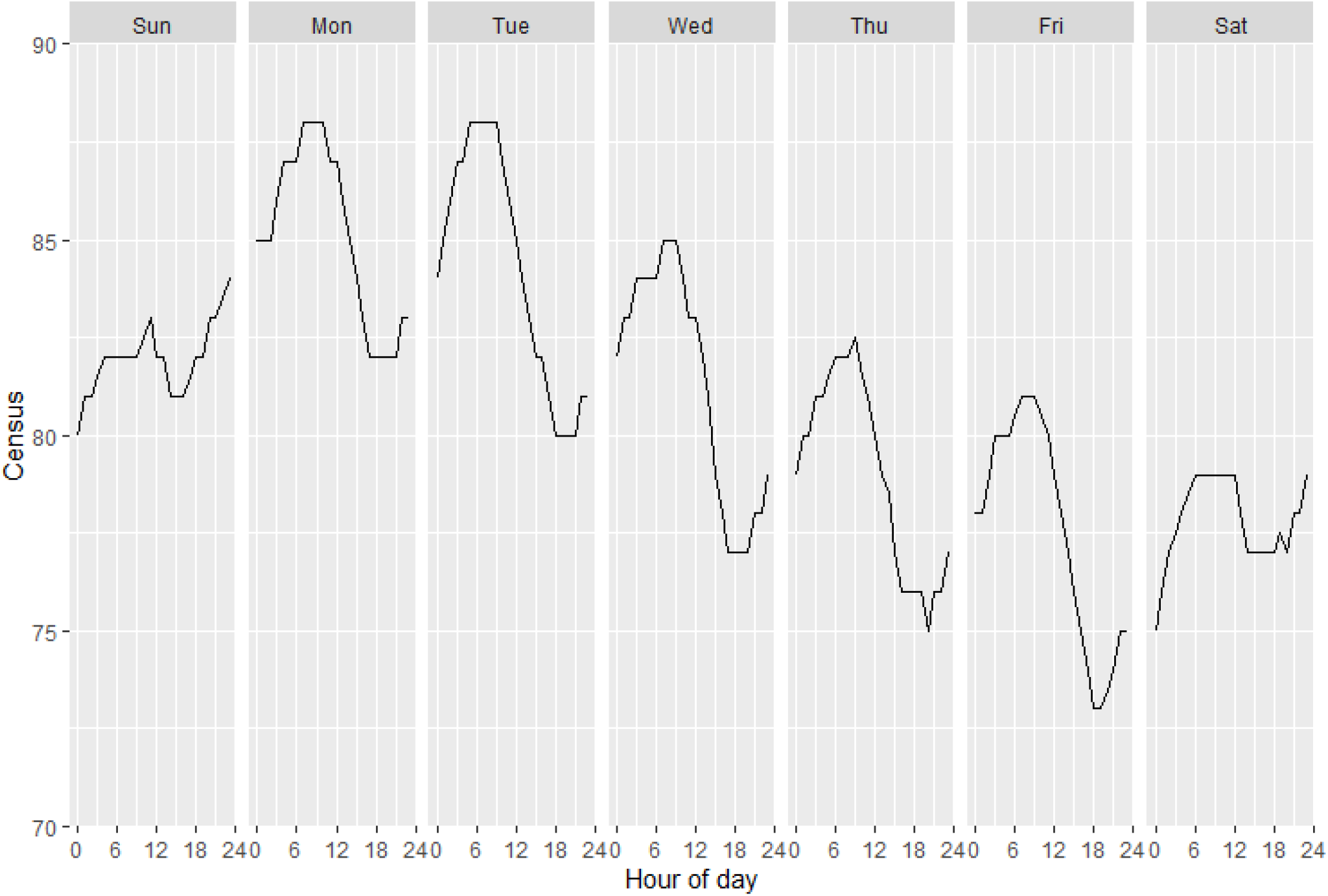
Median census by hour of day and day of week Legend: Data ranged from Sunday, January, 2016 to Saturday, February, 2019, excluding holidays.

**Figure 1C.**
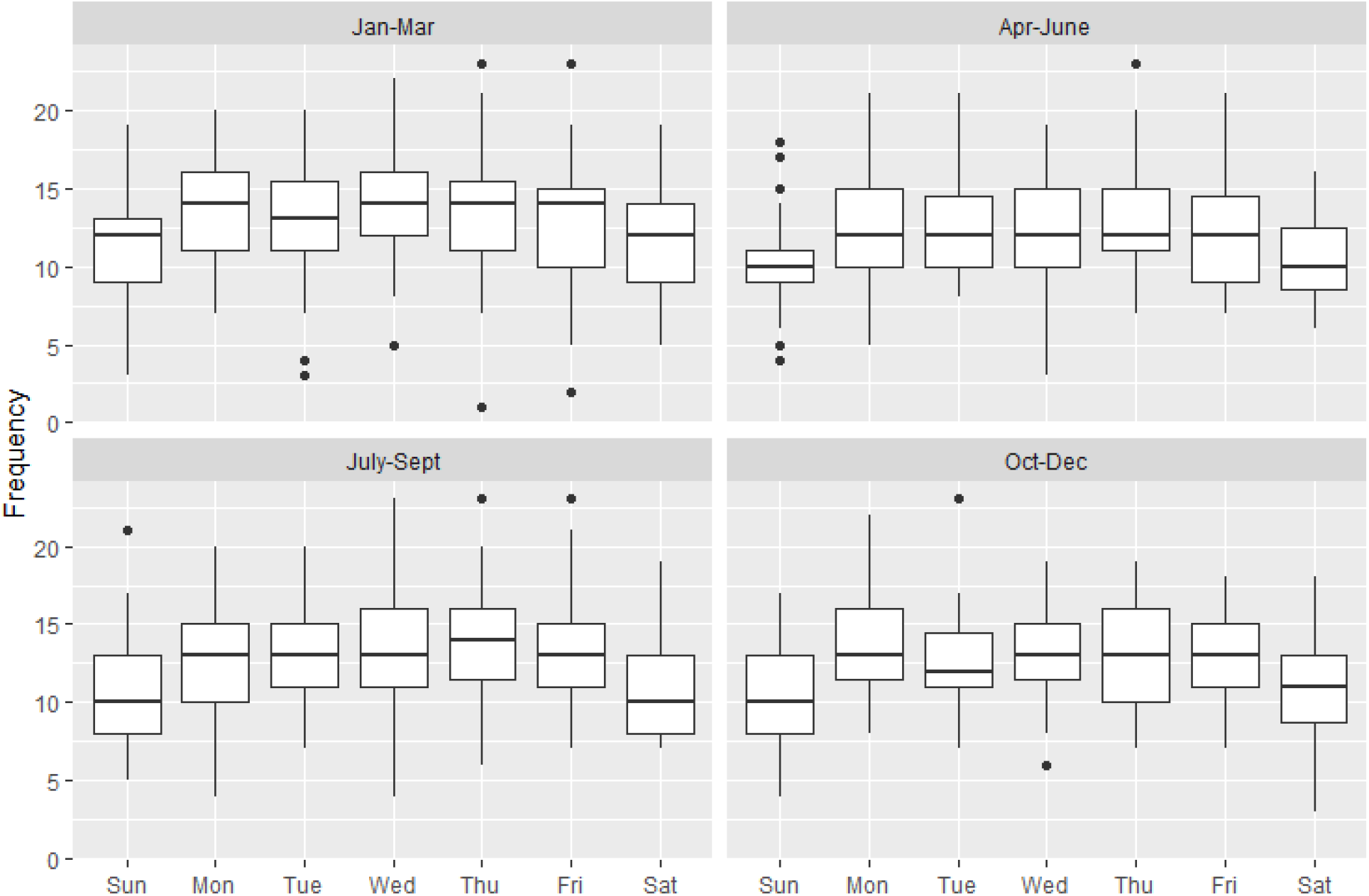
Distribution of daily frequency of internal medicine consults Legend: Data spans from January, 2016 to December, 2018. For the ED consults data, box-whisker plot is used to show an indication of how the daily frequency of ED consults are spread out. The lower and upper bounds of the box represent the 25^th^ percentile and the 75^th^ percentile of the frequencies, respectively, while the horizontal line inside the box represents the median of the data. The range that the box covers (25^th^ percentile to 75^th^ percentile) is defined as interquartile range (IQR), and the lower and upper ends of the whiskers represent the “minimum” (i.e., 25^th^ percentile – 1.5*IQR) and “maximum” (i.e., 75^th^ percentile + 1.5*IQR) observation in the sample, respectively. The black dots are outliers.

**Figure 1D.**
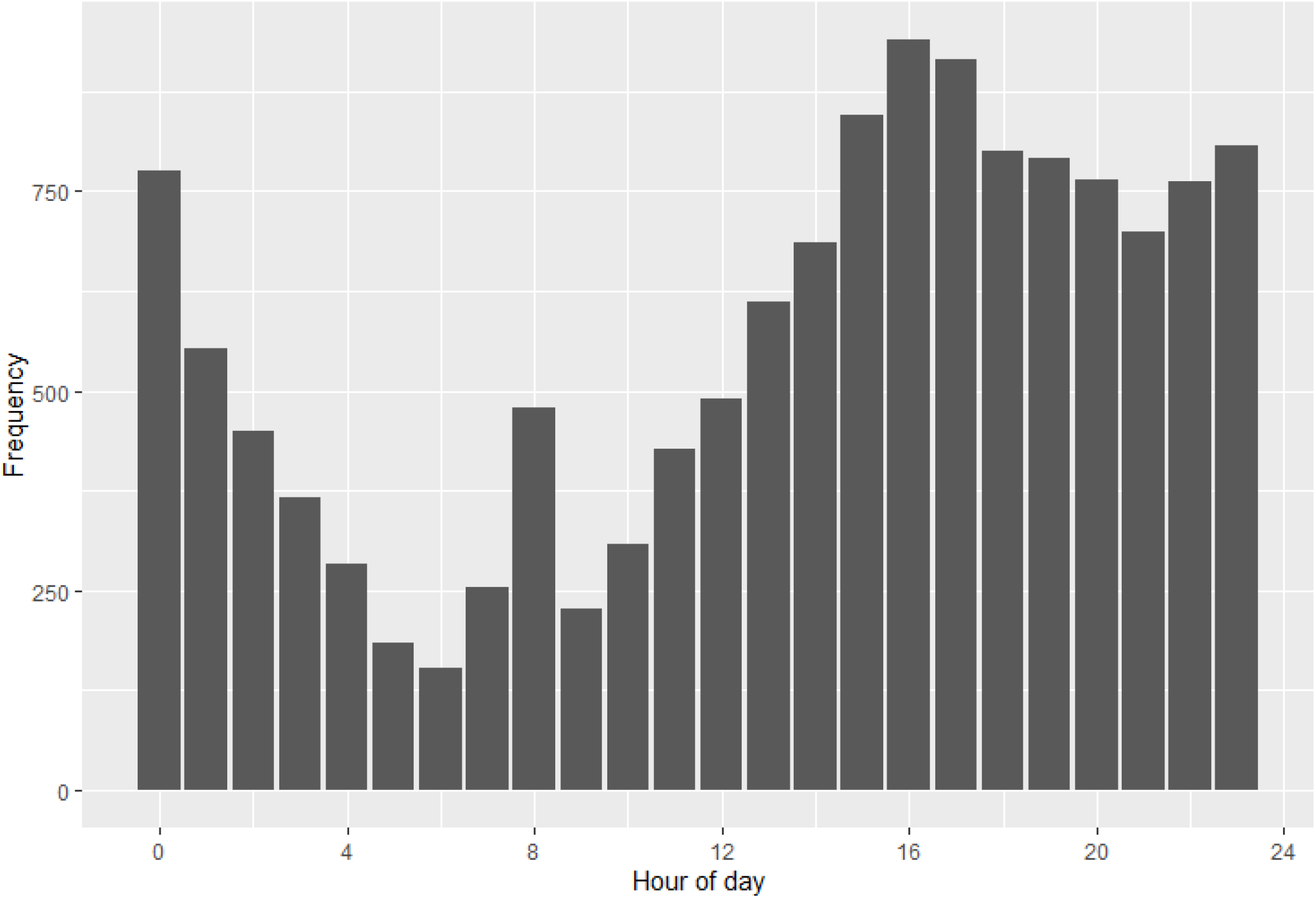
Frequency of ED consults by hour of request (actual or imputed) Legend: Data ranged from January, 2016 to December, 2018. There is an apparent “spike” at 8:00 AM because consultations after 6:00 AM are generally held-over until the new resident arrives at 8:00 AM.

In the current staffing model, resident utilization varied across the three scenarios (Figure 2). Across all three scenarios of various levels of staffing, weekday resident utilization was generally highest between 8:00 AM and 2:00 PM and lowest between 1:00 AM and 8:00 AM (Figure 2). In Scenario 1, resident utilization generally never exceeded 100% utilization. In contrast, in Scenario 2 and Scenario 3 it was common for resident utilization to approach and exceed 100% utilization particularly during the daytime (Figure 2B, Figure 2C). In the sensitivity analysis where we assumed patient care tasks took 50% longer, resident utilization in Scenario 1 began to approach and exceed 100%, and resident utilization in Scenarios 2 and 3 was consistently above 100% (Appendix 2).

**Figure 2A.**
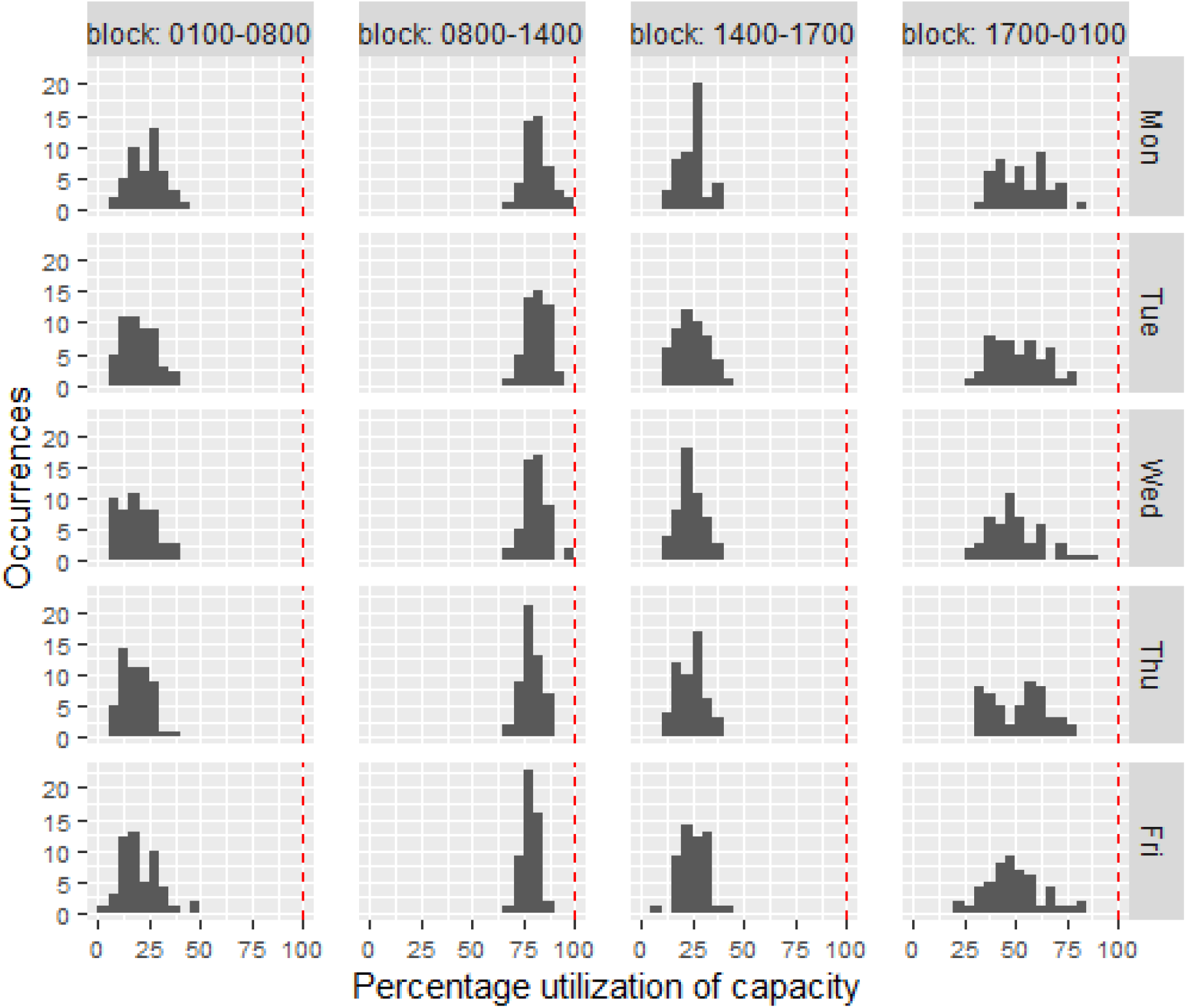
Weekday utilization Scenario 1 Legend: Resident utilization whereby each panel in the utilization histogram graph contains 52 data points representing the 52 weeks of the year. The red dashed line marks the 100% utilization. Scenario 1 assumed all residents were present apart from those who were post-call.

**Figure 2B.**
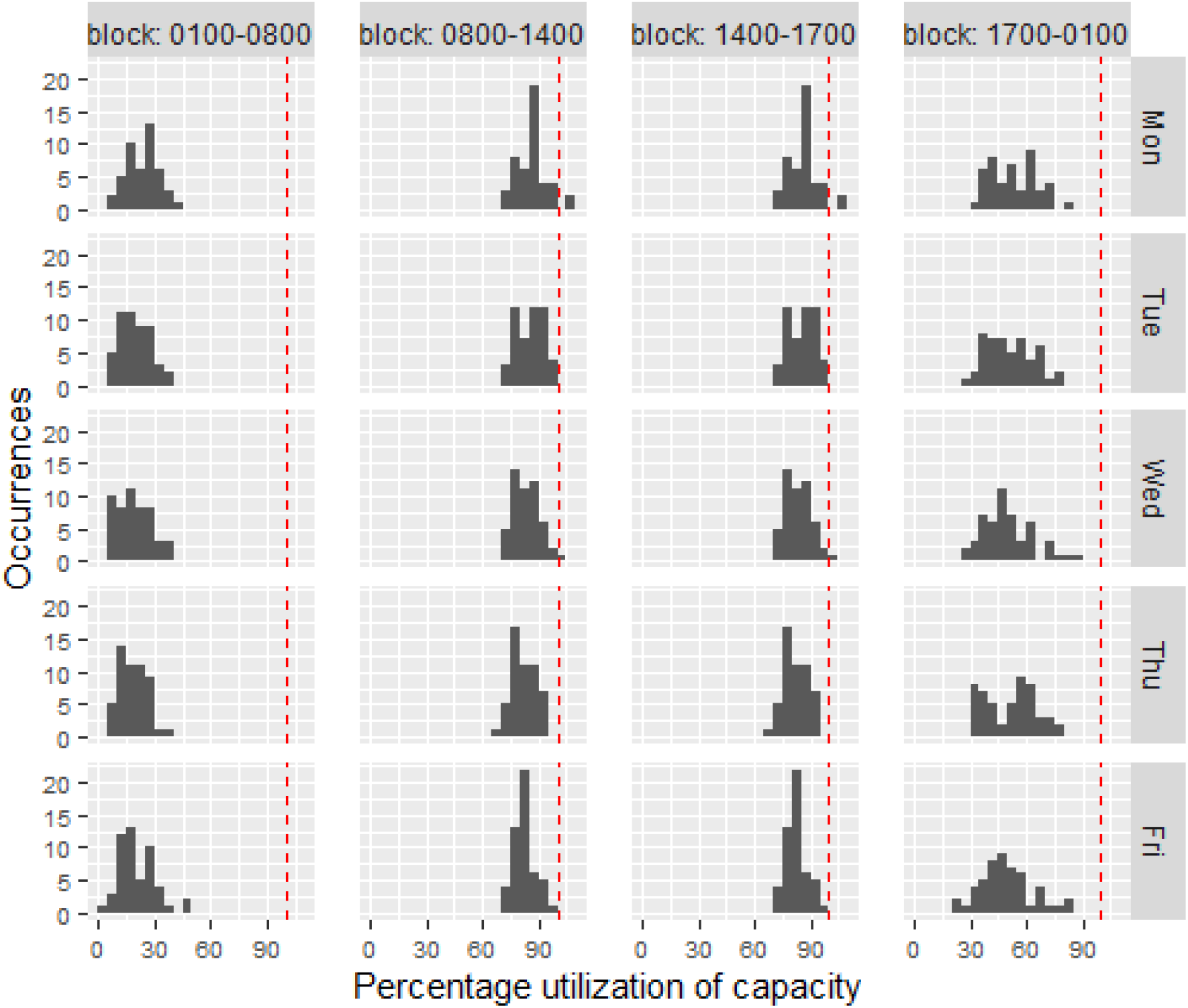
Weekday utilization Scenario 2 Legend: Resident utilization whereby each panel in the utilization histogram graph contains 52 data points representing the 52 weeks of the year. The red dashed line marks the 100% utilization. Scenario 2 assumed one resident was away on vacation each week and also accounted for a resident being post-call.

**Figure 2C.**
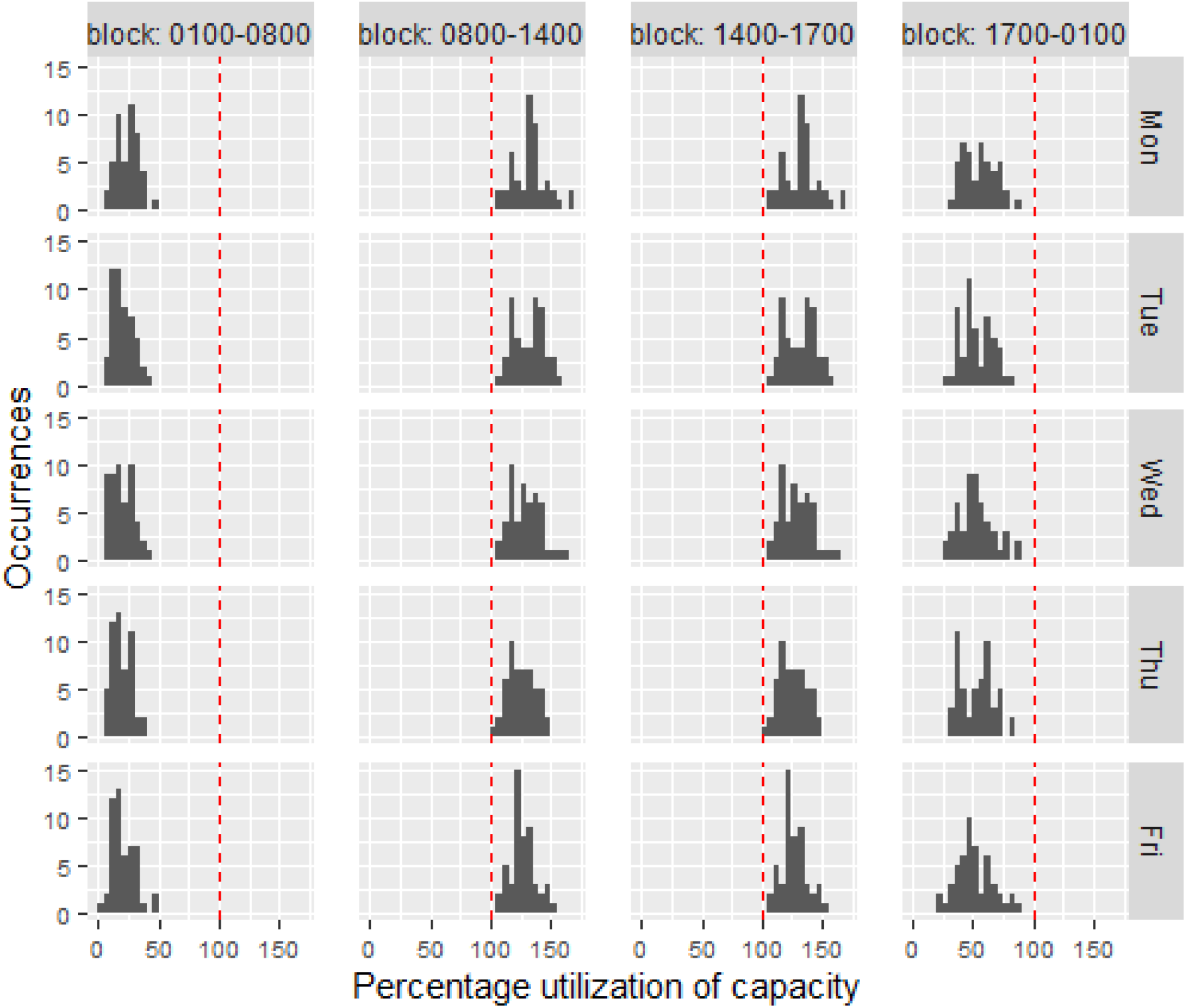
Weekday utilization Scenario 3 Legend: Resident utilization whereby each panel in the utilization histogram graph contains 52 data points representing the 52 weeks of the year. The red dashed line marks the 100% utilization. Scenario 3 assumed one resident was away on vacation and another resident was away sick, and also accounted for a resident being post-call.

For weekend resident utilization, results were consistent across all three scenarios because resident staffing is consistent on the weekends, regardless of any residents being on vacation or away sick. During the weekends, resident utilization was highest from 8:00 AM to 2:00 PM, during which it was consistently close to or reaching 100%; it was lowest from 1:00 AM to 8:00 AM (Figure 2D). In the sensitivity analysis similar results were observed, though 100% utilization commonly occurred during the 2:00 PM to 1:00 AM block.

**Figure 2D.**
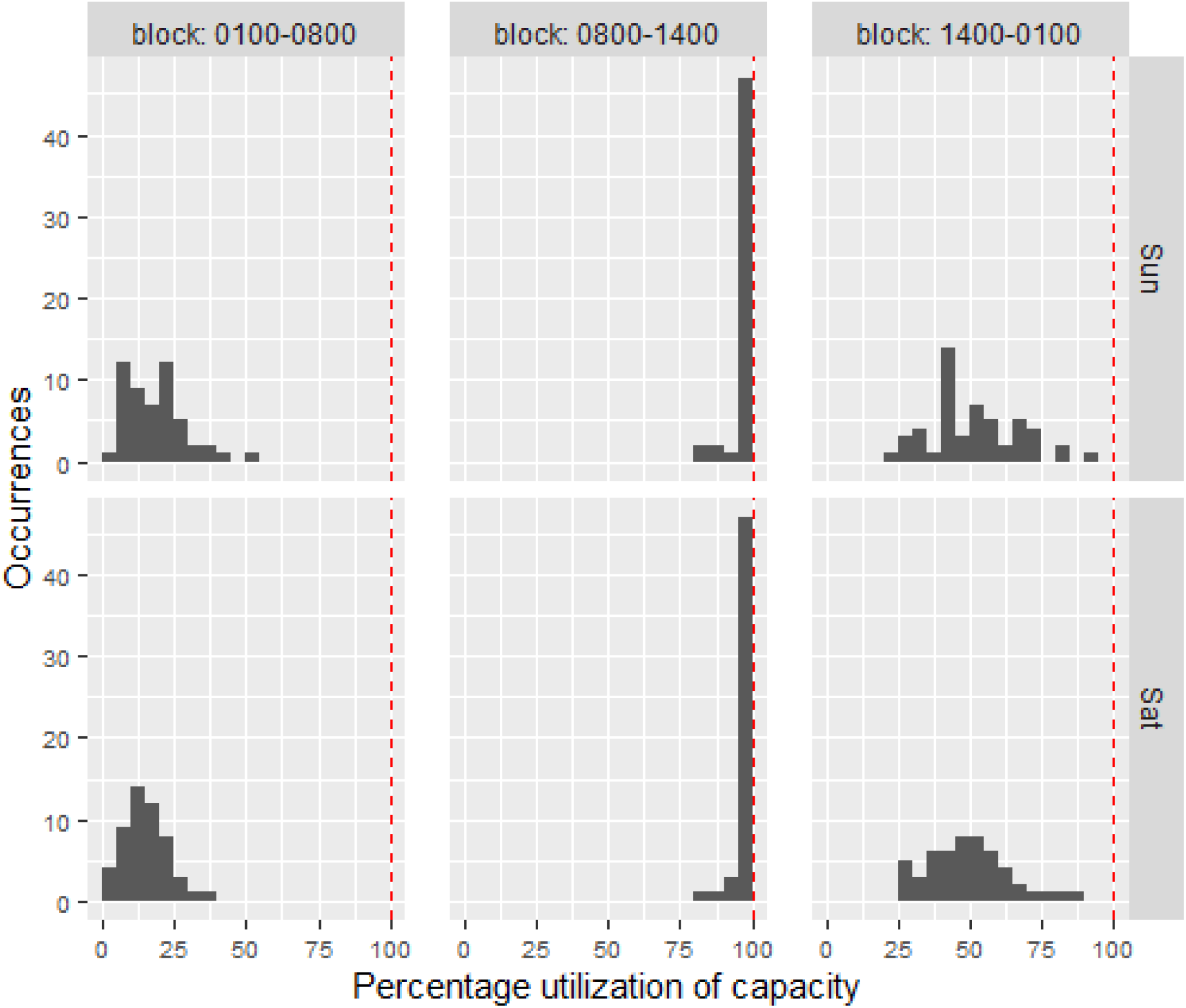
Weekend utilization across all scenarios Legend: Resident utilization whereby each panel in the utilization histogram graph contains 52 data points representing the 52 weeks of the year. The red dashed line marks the 100% utilization.

## INTERPRETATION

### Main Findings

In this single centre study, we were able to model patient care tasks against resident supply to understand how supply and demand changes over the course of a given day and under various levels of resident absenteeism. Doing so allowed us to identify periods of mismatch which provides data to inform how the schedule might be augmented to improve efficiency. Furthermore, our results highlight the vulnerability of the current scheduling model when residents are away.

The model can also be used to estimate whether a given number of residents would result in acceptable utilizations, under scenarios where the volume of patient care tasks or the durations of patient care tasks have changed. For example, during the first wave of COronaVirus Disease 2019 (COVID-19) many hospitals observed increased volumes of admitted patients and increased time required to perform patient care tasks due to requirements for personal protective equipment.^7,8^ These two factors would affect the demand on the ward, and given input data quantifying the change, the model could be used to estimate whether a proposed modification to the resident schedule would provide sufficient supply to satisfy demand at acceptable utilization rates. Determining the ideal utilization of healthcare team members will vary depending on whether a hospital’s goal is to have members working at maximum capacity (i.e., 100%) or working marginally below maximum (e.g., 80%) to allow for potential surges in workload.

### Comparison with Other Studies

While there are few empirical studies that have been published in this area, our results mirror the experience of academic general internists who work on the inpatient setting. For example, it is well known among physicians working on a general internal medicine service at a teaching hospital that the current scheduling approach is highly vulnerable to resident absenteeism and it is common to have only one resident present each day after vacations, illness, and teaching sessions are accounted for. We were able to demonstrate this by modeling the supply and demand across various scenarios of resident absenteeism to empirically quantify how this leads to overutilization (i.e., demand exceeds supply). Notably, the different scenarios we modeled did not affect supply and demand on the weekend, though this reflects the fact that, regardless of the number of residents scheduled for a general internal medicine rotation, the weekend coverage is fixed at one resident per day. In contrast, the different scenarios clearly demonstrated how weekday demand exceeds supply with increasing absenteeism. This observation likely explains why many teaching hospitals have hired physician-extenders such as physician assistants or nurse practitioners to provide consistent levels of staffing. Other hospitals have also shifted their scheduling approach to a night float system, which again serves to provide more consistent daytime staffing of residents because the residents are not post-call and therefore not absent from the clinical ward during the daytime. Of course, each approach has important implications spanning from financial in the case of hiring physician extenders, to resident fatigue and work-life balance in the case of the night float system.

### Limitations

There are a number of important limitations to our study. First, it was single centre and thus our results may not generalize to other centres. Specifically, our results will not generalize to community-based hospitals that do not have medical trainees, nor will it necessarily generalize to hospitals that have a hybrid approach of teams cared for by physician-extenders (e.g., nurse practitioners, physician assistants) rather than medical trainees. Second, while we attempted to account for all of the various patient care tasks, there were others that we did not include due to a lack of data for those tasks (i.e., bedside procedures, hand-over, updating the medical sign-out list, calling for consultations). Third, for tasks with durations no recorded in the EMR, we estimated the amount of time for each task included in our model based on input from a relatively small and convenience sample of trainees and staff physicians, but our model does not account for the fact that time spent on any given task likely varies between residents. To help account for this, we conducted a sensitivity analysis in which we assumed tasks took 50% longer. This sensitivity analysis demonstrated that Scenario 2 and Scenario 3 were particularly affected by a 50% increase in task duration, as demonstrated by utilizations that consistently exceeded 100%. Another limitation of our study is that our model did not account for how “hands-on” the attending physician was and how this might vary across different levels of resident absenteeism to help balance supply and demand.

## Conclusions

While our results were drawn from a single institution, we anticipate that many other hospitals also schedule the work hours of physicians based on historical scheduling practices. Scheduling physician work hours to better align with local patient demand may represent a more efficient and patient-centred approach. An added benefit of having a mechanism to model supply and demand is the ability to recalibrate scheduling as patient demand changes.

## Data Availability

Data from this study are not available.

## APPENDIX 1 Data sources and statistical analysis

– GIM census, transfer and discharge data was available from 2016 to 2019
– ED consult data was available from January, 2016 to December, 2018
– Paging data were available from February 2018 to February 2019
– Since paging data were only available from February 2018 to February 2019, we estimated resident utilization using this one-year period for all data sources (except cardiac arrests).
– Cardiac arrest data were only available from 2013 until June 2017, so we used the one-year period from February 2016 to February 2017, and aligned it with the other data sources.
– For each day, the number of patients to round on is set equal to (8:00 AM census - patients discharged during 8:00 AM to 6:00 PM) X (percentage of patients to round on), every day of the week. “Percentage of patients to round on” is set to 90% for weekdays, 75% for weekends. For each day in Scenario 1, rounding is allocated to the first period (which starts at 8:00 AM), until the resident hours in that period are fully utilized, and then remaining rounds are allocated to the following period. In Scenarios 2 and 3, rounding is allocated between the first two periods in a manner such that the resident utilizations in the two periods are equal.
– When a cardiac arrest occurs, we assume that the number of residents who are occupied by the cardiac arrest is 3 or the total number of residents on the responsible team, whichever is less.
– ED consult durations for senior residents, junior medicine residents, and junior non-medicine residents (Appendix Table 1) are used to compute a weighted average ED consult duration during each period of the week. The weighted average is based on the ratios of junior and senior residents working on the CTU. When one or more junior residents are working, we assume that one is medicine and the rest are non-medicine.

**Appendix Table 1:** Durations used for each patient care task in the modeling of demand. Cardiac arrest durations were estimated using the median duration in the hospital’s database. Durations for all other tasks were estimated from interviews with staff and residents.

| **Task** | **Duration (minutes)** | **Sensitivity analysis** |
| --- | --- | --- |
| ED consult (Senior Resident, per consult) | 60 | 90 |
| ED consult (Junior Medicine Resident, per consult) | 90 | 135 |
| ED consult (Junior Non-Medicine Resident, per consult) | 150 | 225 |
| ICU transfer (per patient) | 60 | 90 |
| Page (per page) | 5 | 7.5 |
| Cardiac arrest (per instance) | 22 | 33 |
| Cancelled cardiac arrest (per instance) | 2 | 3 |
| Rounding (per patient) | 20 | 30 |
| Discharge (per patient) | 30 | 45 |
| Non-patient-related ward tasks (per resident per day) | 120 | 180 |

**Appendix 2 Figure 1.**
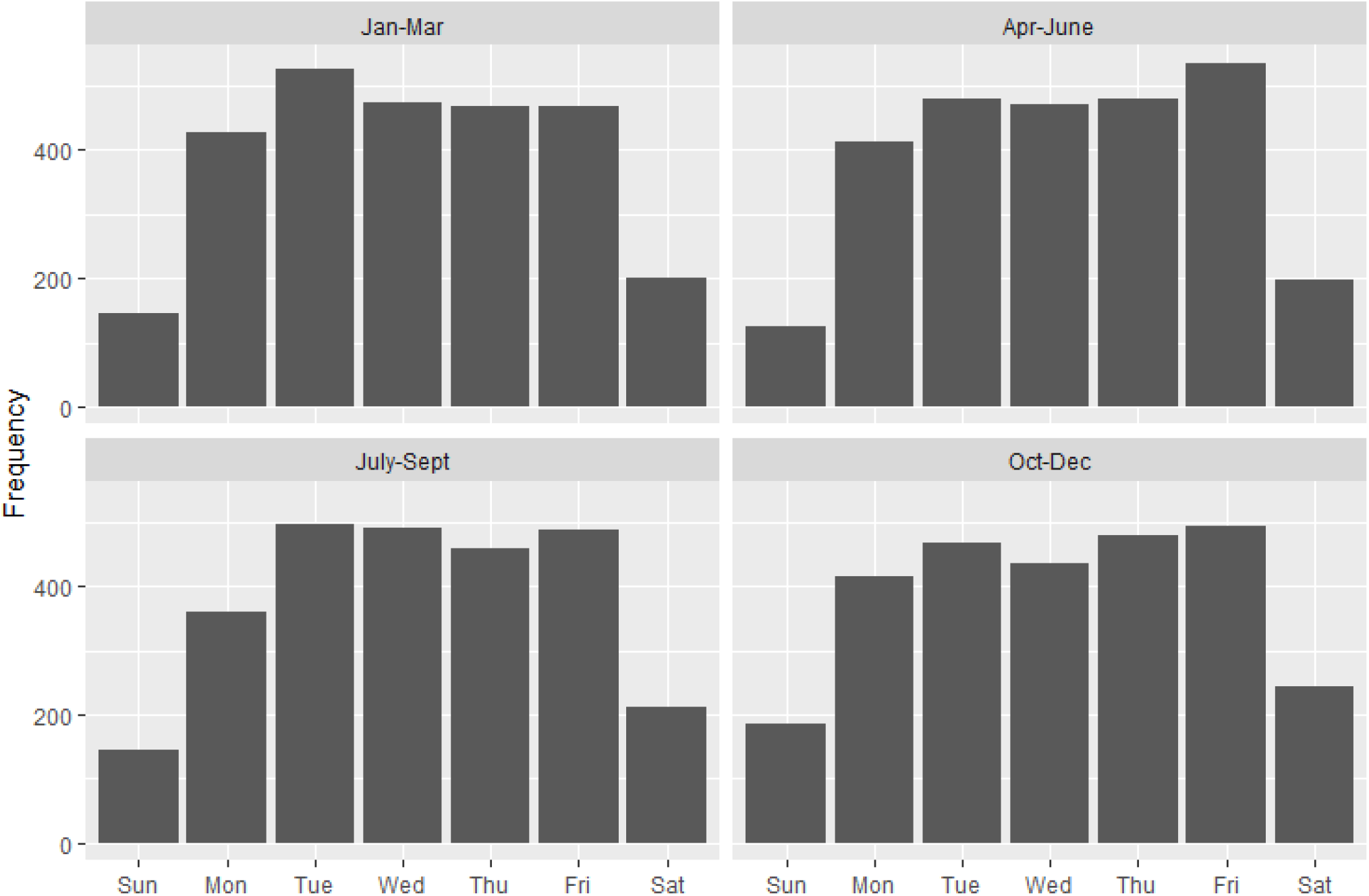
Frequency of hospital discharges

**Appendix 2 Figure 2.**
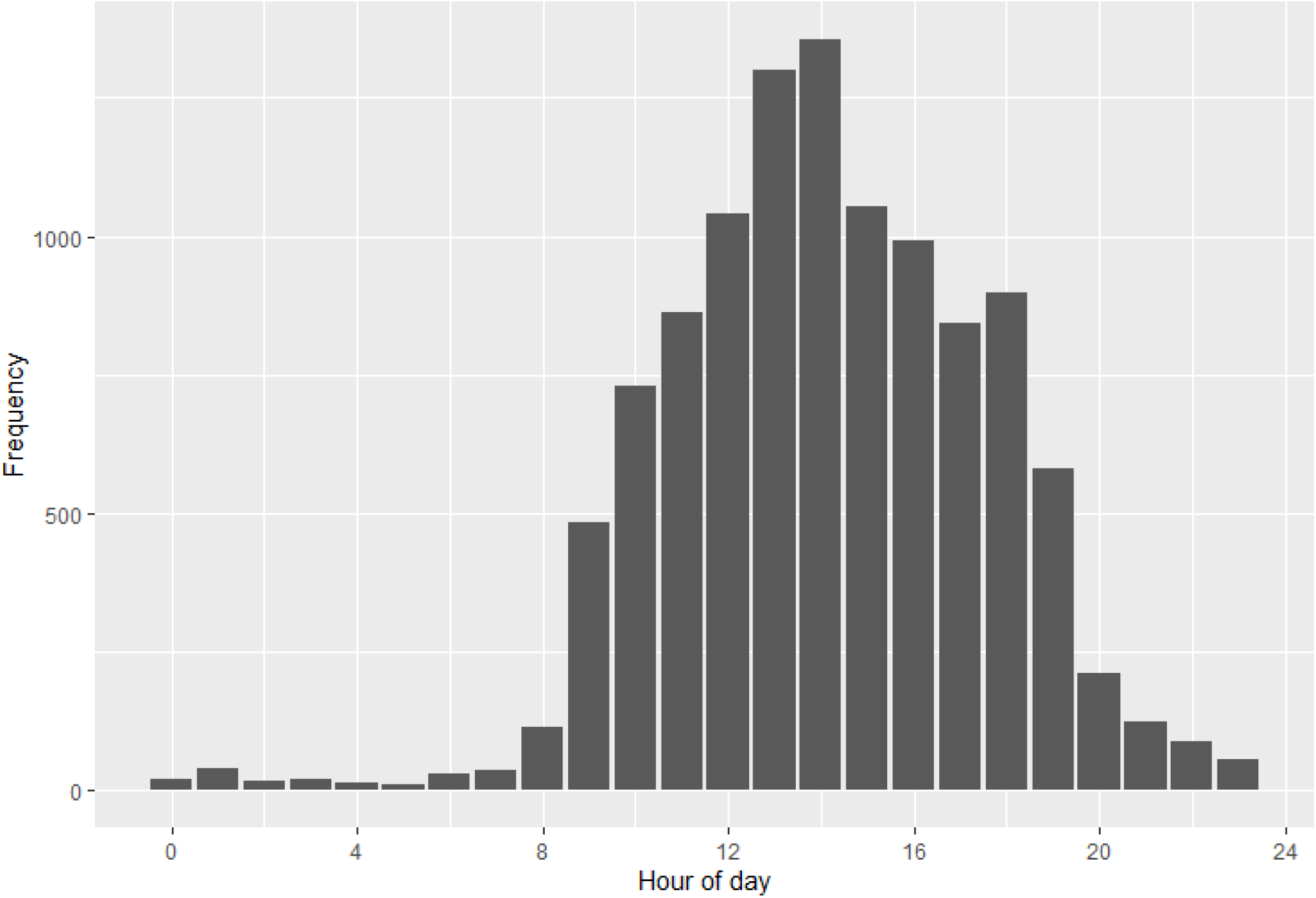
Hospital discharges by hour of the day

**Appendix 2 Figure 3.**
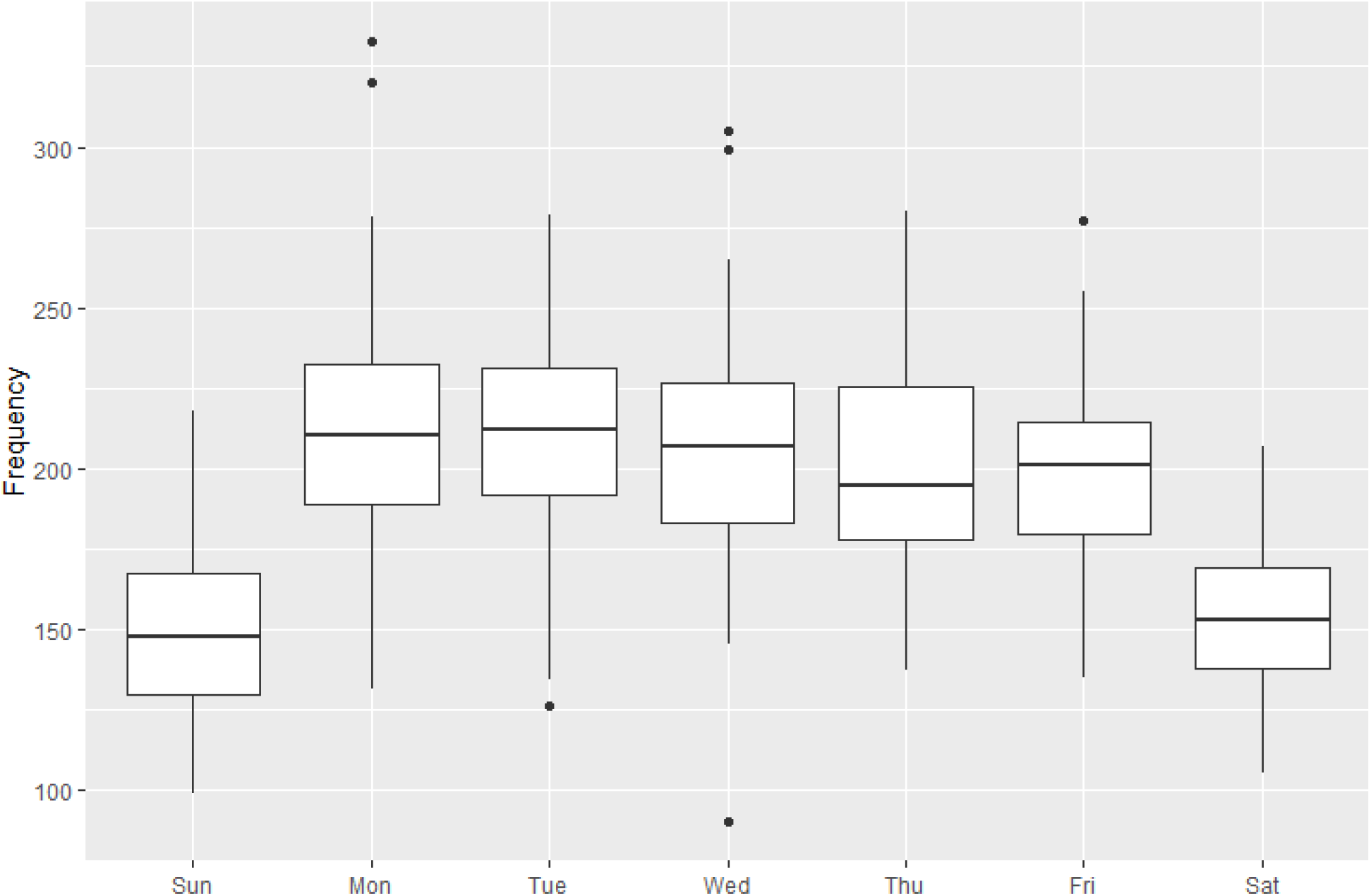
Distribution of pages by day of the week Legend: For the frequency of pages data, box-whisker plot is used to show an indication of how the data values are spread out by each day of the week. The lower and upper bounds of the box represent the 25^th^ percentile and the 75^th^ percentile of the frequencies, respectively, while the horizontal line inside the box represents the median of the data. The range that the box covers (25^th^ percentile to 75^th^ percentile) is defined as interquartile range (IQR), and the lower and upper ends of the whiskers represent the “minimum” (i.e., 25^th^ percentile – 1.5*IQR) and “maximum” (i.e., 75^th^ percentile + 1.5*IQR) observation in the sample, respectively. The black dots are outliers.

**Appendix 2 Figure 4A.**
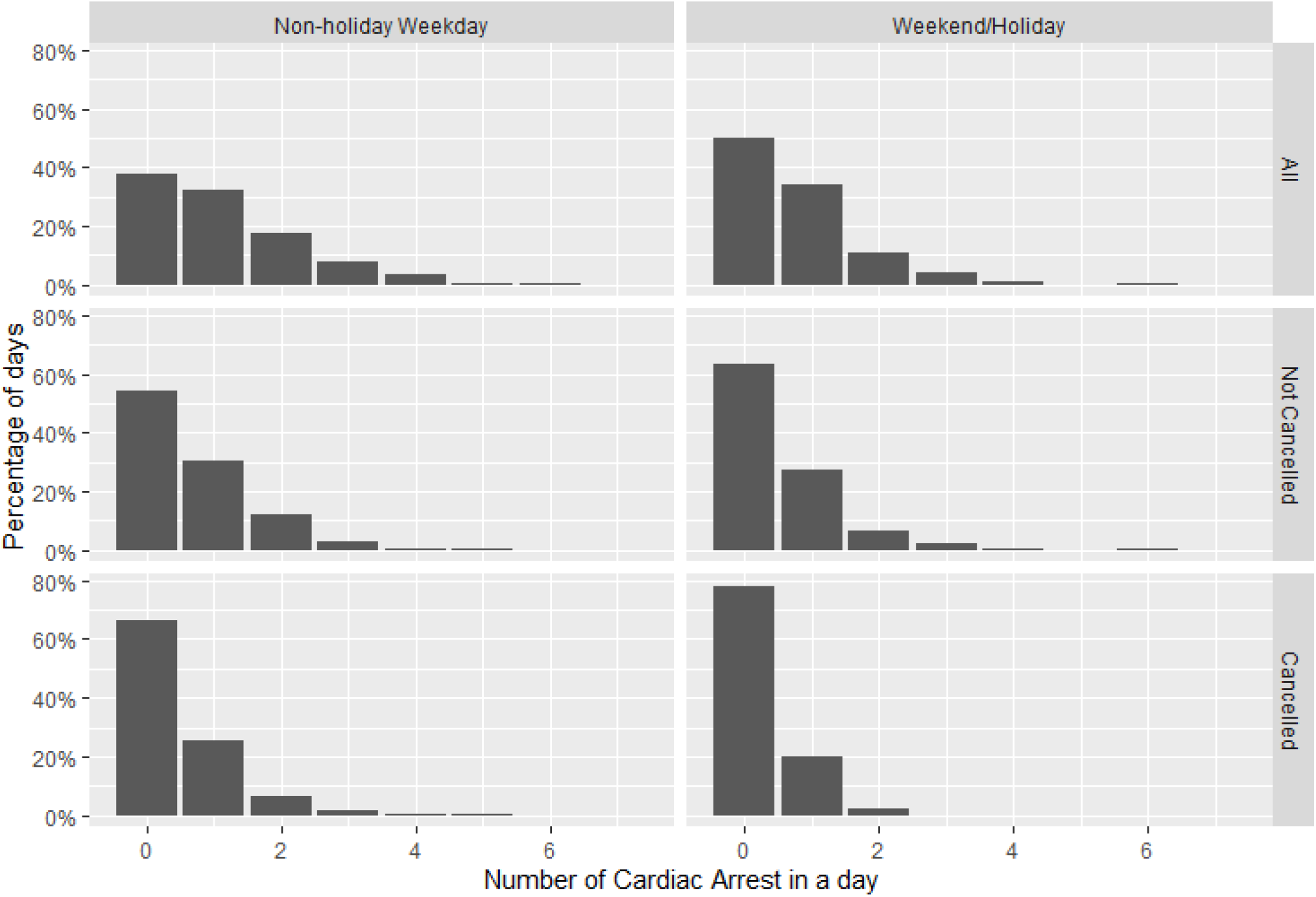
Frequency of cardiac arrests

**Appendix 2 Figure 4B.**
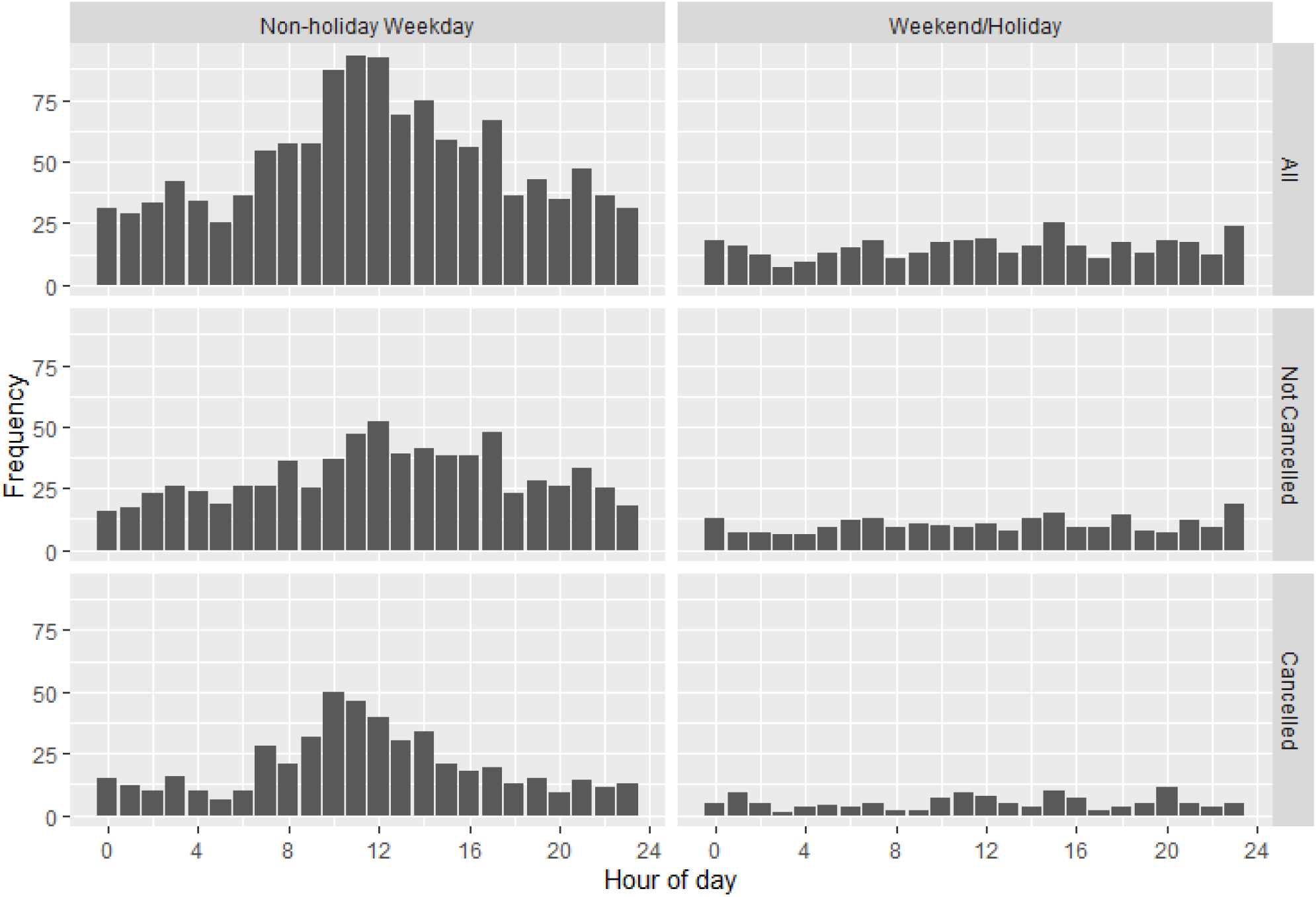
Frequency of cardiac arrests by hour of the day Legend. Figure 4a shows the percentage of days that have either 0 or more cardiac arrests per day. Figure 4b displays daily frequency of cardiac arrests by hour of the day.

Appendix 2 Figure 5. Results of sensitivity analysis in which patient tasks were assumed to take 50% longer than in the original model

**Appendix 2 Figure 5A.**
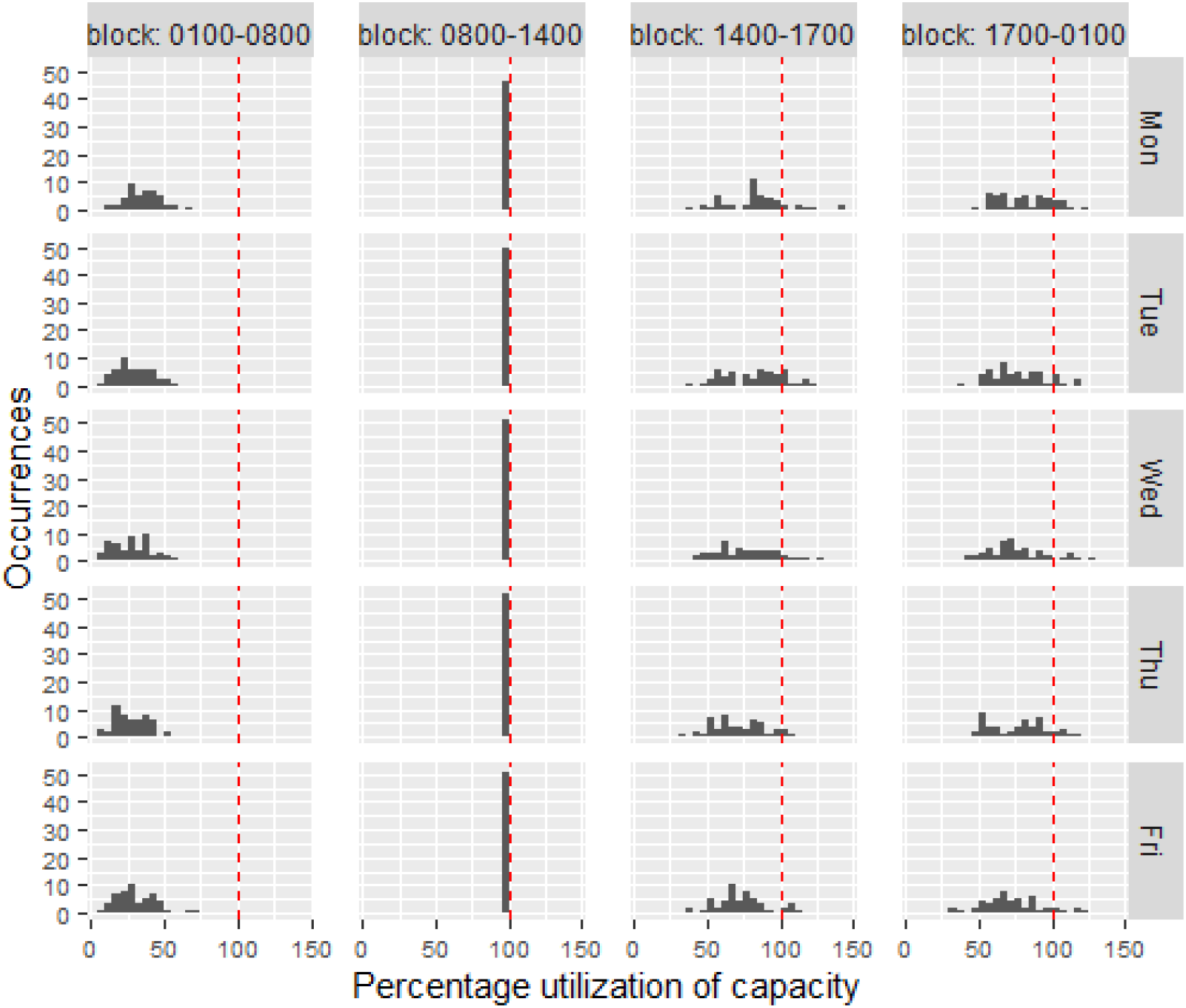
Weekday utilization Scenario 1 sensitivity analysis Legend: Resident utilization whereby each panel in the utilization histogram graph contains 52 data points representing the 52 weeks of the year. The red dashed line marks the 100% utilization.

**Appendix 2 Figure 5B.**
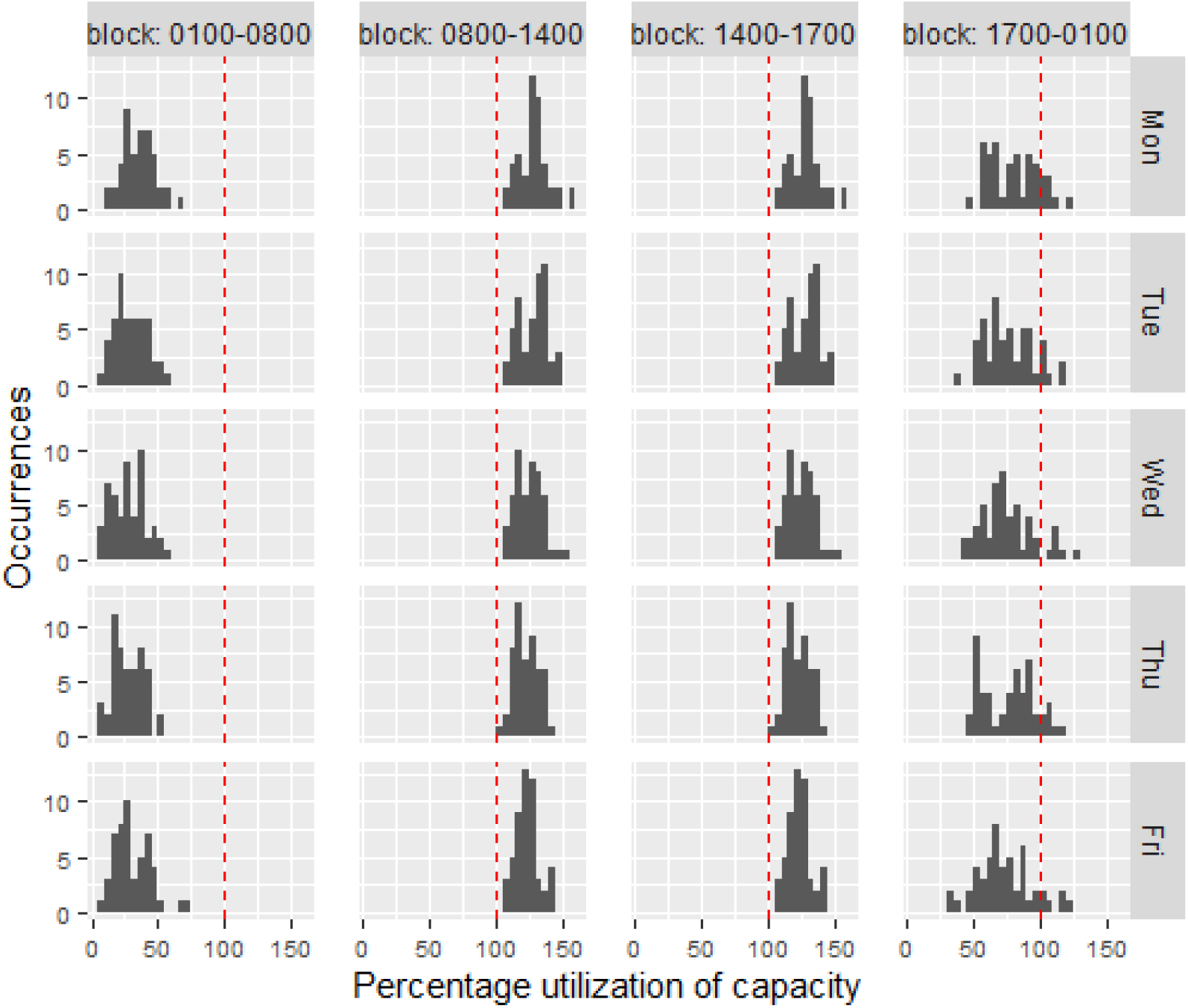
Weekday utilization Scenario 2 sensitivity analysis Legend: Resident utilization whereby each panel in the utilization histogram graph contains 52 data points representing the 52 weeks of the year. The red dashed line marks the 100% utilization.

**Appendix 2 Figure 5C.**
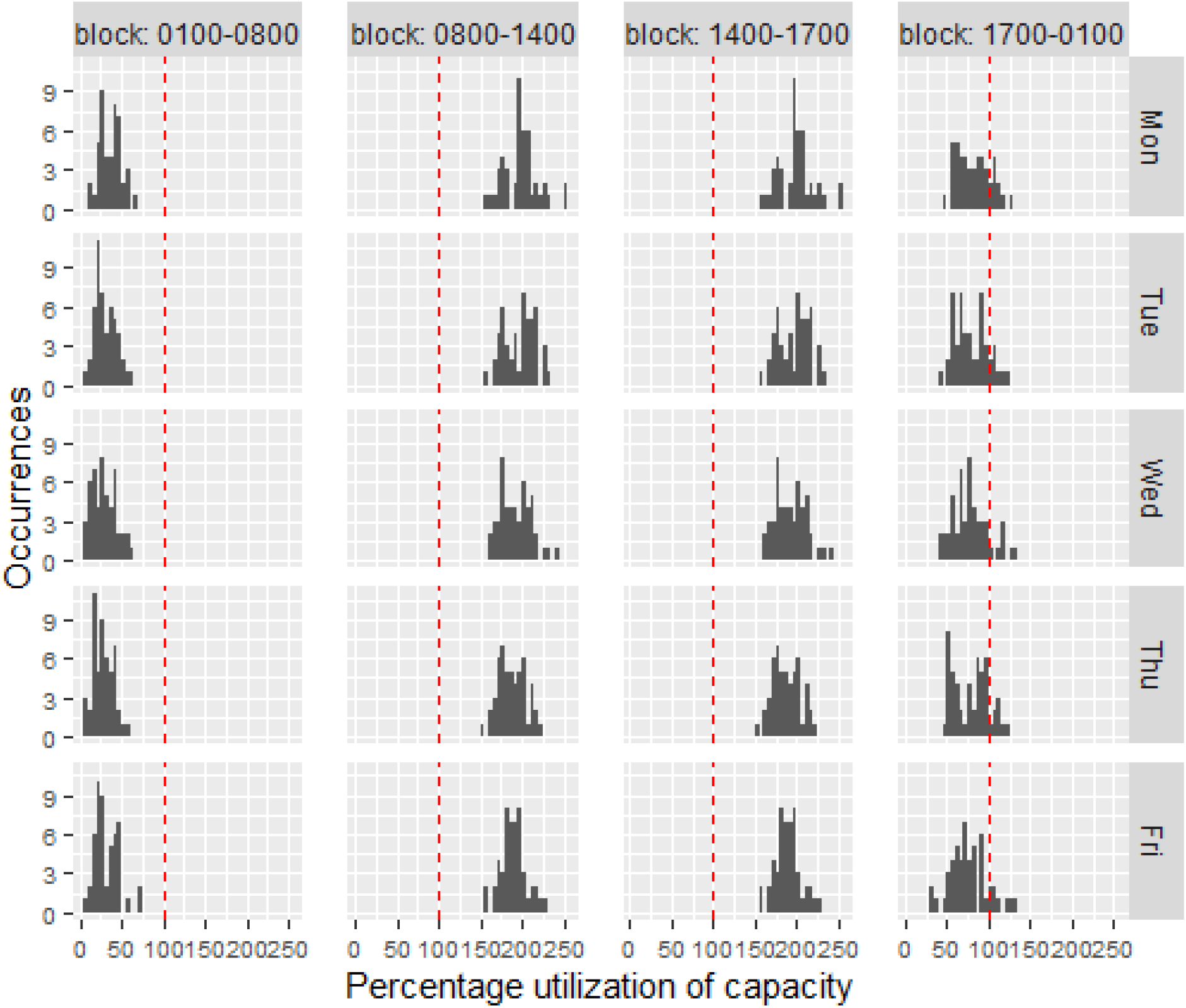
Weekday utilization Scenario 3 sensitivity analysis Legend: Resident utilization whereby each panel in the utilization histogram graph contains 52 data points representing the 52 weeks of the year. The red dashed line marks the 100% utilization.

**Appendix 2 Figure 5D.**
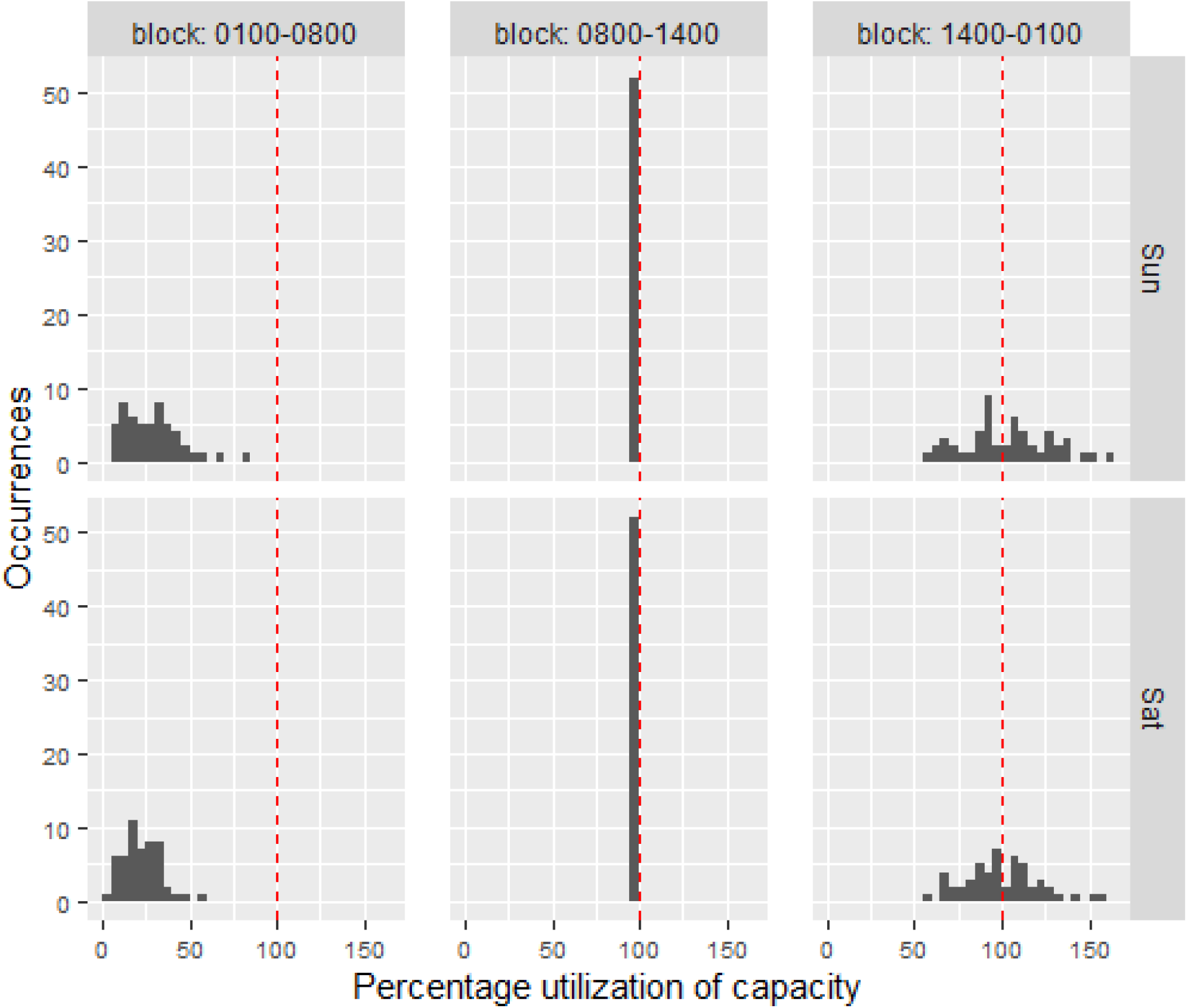
Weekend utilization across all scenarios sensitivity analysis Legend: Resident utilization whereby each cell in the utilization histogram graph contains 52 data points representing the 52 weeks of the year. The red dashed line marks the 100% utilization.

## Notes

**Conflicts of interest**: Dr. Fralick reports no relevant conflicts of interest.

### Competing Interest Statement

The authors have declared no competing interest.

### Funding Statement

This study was funded by the Li Ka Shing Foundation.

### Author Declarations

This study was approved by the St. Michael's Hospital Research Ethics Board.

## REFERENCES

1. Weinberger SE, Pereira AG, Iobst WF, Mechaber AJ, Bronze MS. Annals of Internal Medicine Academia and the Profession Competency-Based Education and Training in Internal Medicine. Ann Intern Med. 2010.

2. Frank JR, Snell LS, Cate O Ten, et al. Competency-based medical education: theory to practice. Med Teach. 2010;32(8):638–645.

3. Verma AA, Guo Y, Kwan JL, et al. Patient characteristics, resource use and outcomes associated with general internal medicine hospital care: the General Medicine Inpatient Initiative (GEMINI) retrospective cohort study. CMAJopen. 2017;5(4):E842–E849.

4. McMahon GT, Katz JT, Thorndike ME, Levy BD, Loscalzo J. Evaluation of a Redesign Initiative in an Internal-Medicine Residency. N Engl J Med. 2010;362(14):1304–1311.

5. Sharma A, Lo V, Lapointe-Shaw L, Soong C, Wu PE, Wu RC. A time-motion study of residents and medical students performing patient discharges from general internal medicine wards: a disjointed, interrupted process. Intern Emerg Med. 2017;12(6):789–798.

6. Michtalik HJ, Yeh HC, Pronovost PJ, Brotman DJ. Impact of attending physician workload on patient care: A survey of hospitalists. JAMA Intern Med. 2013;173(5):375–377.

7. Docherty AB, Harrison EM, Green CA, et al. Features of 16, 749 hospitalised UK patients with COVID-19 using the ISARIC WHO Clinical Characterisation Protocol. 2020.

8. Jeffery MM, D’Onofrio G, Paek H, et al. Trends in Emergency Department Visits and Hospital Admissions in Health Care Systems in 5 States in the First Months of the COVID-19 Pandemic in the US. JAMA Intern Med. 2020;06519:1–6.

